# Impact of brain frailty on complications and outcomes in acute ischemic stroke

**DOI:** 10.1101/2024.11.01.24316618

**Authors:** XiaoQing Cheng, Wu Cai, ChuYan Li, Xi Shen, JiaNan Li, LiJun Huang, JinJing Tang, HuiMin Pang, BaiYan Luo, Ya Liu, QianChen, Lulu Xiao, WuSheng Zhu, ZeHong Cao, XiaoYu Liu, LongJiang Zhang, XinDao Yin, ZhiQiang Zhang, Feng Shi, Wei Xing, GuangMing Lu

## Abstract

**BACKGROUND:** This study aimed to determine the association of brain frailty with acute complications and prognosis in patients with acute ischemic stroke (AIS) due to occlusion of large vessels in the anterior circulation, and to further assess its predictive value.

**METHODS AND RESULTS:** This multicenter, retrospective study included patients with AIS due to large vessel occlusion in the anterior circulation. All patients underwent MRI within seven days of stroke onset, measuring subcortical and cortical atrophy and leukoaraiosis as indicators of brain frailty.The study included 1,090 patients with a median age of 64 (interquartile range, 55–73) years and a median National Institutes of Health Stroke Scale (NIHSS) score of 9 (interquartile range, 4.5–15). Multivariable logistic regression analysis showed that independent risk factors for the unfavorable clinical outcome included: NIHSS score (OR, 1.17; 95% CI, 1.13–1.22), blood glucose (OR, 1.15; 95% CI, 1.04–1.26), infarct volume (OR, 1.32; 95% CI, 1.15–1.52), subcortical atrophy (OR, 1.27; 95% CI, 1.18–1.37), severe cortical atrophy (OR, 5.46; 95% CI, 1.71–17.45), and severe leukoaraiosis (OR, 4.68; 95% CI, 1.93–11.31). However, brain frailty was not significantly associated with AIS complications (malignant cerebral edema, parenchymal hemorrhage). Including brain atrophy indicators in the model significantly improved its unfavorable clinical outcome predictive power (AUC increased from 0.762 to 0.822; *p* < 0.001). The results remained stable in subgroup analyses across treatment modalities.

**CONCLUSIONS:** Brain frailty was significantly associated with the unfavorable clinical outcome but not with acute complications. Brain frailty indicators contributed to the predictive efficacy, regardless of treatment modality.

## INTRODUCTION

Frailty is a clinically recognizable state of reduced physiological reserves in which older adults are more likely to experience a state of poor homeostasis resolution after a stressful event, increasing the risk of adverse outcomes .^1, 2^ Acute ischemic stroke (AIS) represents a quintessential stressor event, with up to 28% of patients with stroke having pre-existing frailty on hospital admission.^3^ The analysis of data from 483,033 participants in the UK Biobank revealed an association between physical frailty and health-related outcomes, which could be mediated by differences in brain structure.^4^ These differences are primarily characterized by hyperintensity on MRI and lower subcortical grey matter volume.^4^ These brain frailty indicators facilitate a more objective assessment than physical signs of frailty.

Evidence suggests indicators of brain frailty are important for stroke prognosis. Post hoc analyses of randomized controlled trials showed that CT-assessed indicators of brain frailty were associated with decreased functional independence in patients with AIS who received intravenous thrombolysis^5^ or endovascular therapy^6^. However, studies using MRI-assessed indicators of cerebral small-vessel disease, such as lacunes, enlarged perivascular spaces, and white matter hyperintensities, indicated that they are not associated with functional outcomes.^7, 8^ Therefore, the choice of indicators might affect findings when assessing the relationship between brain atrophy and prognosis. Furthermore, it is unknown whether brain frailty has a differential effect on acute complications and functional outcome at 90 days.

Various factors influence AIS prognosis. Commonly used indicators to assess stroke complications and functional prognosis include age, National Institutes of Health Stroke Scale (NIHSS) score, infarct volume, blood glucose, and comorbidities.^9–11^ Clinicians combine multiple variables in their clinical decision-making process when treating individual patients. However, few studies have used indicators of brain frailty as an important component in predicting outcomes. Moreover, whether brain frailty enhances models’ predictive power remains unknown.

This multicenter, retrospective study analyzed real-world patients with AIS caused by anterior circulation large-vessel occlusion who received various treatment modalities. The selected indicators of brain frailty included subcortical and cortical atrophy and leukoaraiosis, which can be obtained by simple measurements on CT and MRI scans and are suitable for use in prospective clinical applications. This study aimed to 1) assess the association between brain frailty indicators and AIS complications (malignant cerebral edema, parenchymal hemorrhage) and unfavorable functional outcomes at 90 days and 2) assess the ability of brain frailty indicators to predict outcomes and increase predictive efficacy.

## METHODS

This study was approved by the local ethics committees of all participating institutions (2024DZKY-019-01). The requirement for informed consent was waived by the local ethics committees. We followed the Standards for the Reporting of Diagnostic Accuracy Studies (STARD) guidelines for study execution and reporting (Supplemental Material). Data underpinning the findings of this study can be obtained from the corresponding author upon a reasonable request.

### Study population

This multicenter, retrospective, observational cohort study used data from five centers. We considered consecutive patients presenting with AIS due to large-vessel occlusion in the anterior circulation treated between May 2018 and June 2023. The inclusion criteria were (1) adult patients (aged ≥18 years); (2) underwent MRI within seven days of admission; (3) presence of anterior circulation vessel occlusion (internal carotid or middle cerebral artery M1 or M2); (4) the 90-day modified Rankin Scale (mRS) score was not missing. Patients were excluded if they underwent the MRI beyond seven days or had an intracranial tumor, AIS in the posterior circulation or anterior cerebral artery supply area, severe artifacts, considerable midline shift, and segmentation failure that made it impossible to measure the infarct volume.

### Clinical assessment and outcomes

We retrieved demographic, clinical, laboratory, and imaging data from the electronic medical record, including information on the patient’s age and sex, comorbidities such as hypertension and diabetes mellitus, history of atrial fibrillation and stroke, and the NIHSS score at admission. Laboratory values included blood glucose, glycated hemoglobin, low-density lipoprotein, high-density lipoprotein, and total cholesterol on admission.

The primary outcome was the 90-day functional outcome, assessed using the mRS, obtained by clinical follow-up or a telephone call. The mRS is an ordinal scale used to measure the degree of disability in daily activities. It ranges from 0 (no symptoms) to 6 (death) .^12^ An mRS of 3 to 6 was defined as an unfavorable clinical outcome. The secondary outcomes were malignant cerebral edema and parenchymal hemorrhage. Malignant cerebral edema was defined as a large occupying infarct in the middle cerebral artery compressing the ventricles or causing midline shift of ≥5 mm, consciousness level of ≥1 on item 1a of the NIHSS, and no other causes of neurological deterioration.^13^ Parenchymal hemorrhage was determined based on follow-up imaging and following the European Cooperative Acute Stroke Study II criteria.^14^

### Image acquisition

MRI scans are part of routine clinical care. Since data were collected retrospectively from several clinical facilities, we could not ensure uniform scanner type, scanned area, and scanning protocol. Images were acquired using a 1.5-T or 3-T scanner (GE Discovery MR750 3T, Philips Ingenia 3T, Siemens Skyra 3T or Siemens Avanto 1.5T). The scanning sequences included T2-weighted imaging (T2WI), T1-weighted imaging (T1WI), fluid-enhanced inversion recovery (FLAIR), and diffusion-weighted imaging (DWI) sequences. The parameters of MRI acquisition were detailed in Supplementary Table S1.

### Image analysis

#### Brain frailty assessment

Brain frailty (subcortical and cortical atrophy and leukoaraiosis) was assessed on T1WI, T2WI, or FLAIR sequence. The assessment included the contralateral cerebral hemisphere to avoid confusion due to the acute stroke.

Subcortical atrophy was measured by a senior radiologist with 15 years of experience (***) who were blinded to the clinical information. Subcortical atrophy was assessed by calculating the ratio between the inter-caudate distance and the inner table width.^15,16^ The inter-caudate distance was defined as the minimum distance between the head of the caudate nucleus and the septum lucidum at the level of the foramen of Monroe, multiplied by 2. The distance was measured in the contralateral hemisphere to avoid errors due to infarct-related edema.

Cortical atrophy and leukoaraiosis were assessed by two radiologists with six years of neuroimaging experience (** and ***) who were blinded to the clinical information. Disagreements were resolved by a senior radiologist with fifteen years of experience (***). Cortical atrophy was assessed using the global cortical atrophy scale, which evaluates the degree of cortical atrophy by assessing the sulcus’s width and the gyrus’s volume. The scale ranges from 0 (no atrophy) to 3 (severe atrophy).^17^ Leukoaraiosis was visually assessed for periventricular and deep white matter lesions and a 3-level scale was used to classify whole-brain leukoaraiosis: none or mild (score 0-1), moderate (score 2), and severe (score 3) .^18,19^

#### Infarct volume assessment

An acute infarct segmentation model was established based on the VB-Net structure to automatically segment the whole lesion area on DWI with a b-value of 1,000 s/mm^2^ and with apparent diffusion coefficient map reconstruction images. The automatic segmentation sequence was as follows: 1) Extract the brain tissue from the DWI image to reduce the interference caused by the skull and other parts of the image; 2) normalize the extracted brain tissue so that the image grey scale range will be [0, 1]; 3) use the pre-processed image above as the input to the cascade neural network, and perform step-by-step coarse and fine segmentation of the DWI brain image; 4) obtain the lesion segmentation results based on voxel analysis. Two neuroradiologists with six years of neuroimaging experience proofread the automatic segmentation results (** and ***).

#### Statistical analysis

Categorical variables are presented as counts (percentages) and were compared using the χ^2^ test. Continuous variables are presented as median (interquartile range, IQR) or mean (standard deviation, SD) based on their distribution, evaluated with the Shapiro-Wilk test, and compared using the Mann-Whitney *U* test or *t*-test, as appropriate.

Multivariate logistic regression, adjusted for age, sex, NIHSS score, and infarct volume, helped determine whether brain frailty was associated with acute complications and unfavorable 90-day clinical outcomes. We also performed subgroup analysis after dividing the patients based on the treatment modality received. The area under the receiver operating characteristic (ROC) curve (AUC) was used to assess the ability of individual variables at baseline, including the NIHSS score, infarct volume, baseline glucose, and subcortical atrophy, to predict unfavorable clinical outcomes. The AUC was combined with the multivariate variables to assess whether the brain atrophy indicators could predict unfavorable clinical outcome with a significant potentiating effect. The DeLong test compared the AUC of various variables. Statistical significance was set at *p* < 0.05.

Statistical analysis was performed using R (https://www.r-project.org; The R Foundation) and EmpowerStats (https://www.empowerstats.com;X&Y Solutions, Inc).

## RESULTS

### Baseline patient characteristics

Among the 1,337 consecutively included patients with AIS due to acute large-vessel occlusion in the anterior circulation, 247 were considered ineligible (Figure 1): the MRI examination was performed more than seven days after admission in 92 patients; 5 patients had brain tumors; 39 patients had regional strokes in the vertebral basilar, posterior cerebral, or anterior cerebral artery; 24 patients had severe artifacts; 45 patients were missing the T1WI sequence; 11 patients had extensive midline shift; DWI volume calculation failed in 31 patients.

**Figure 1.**
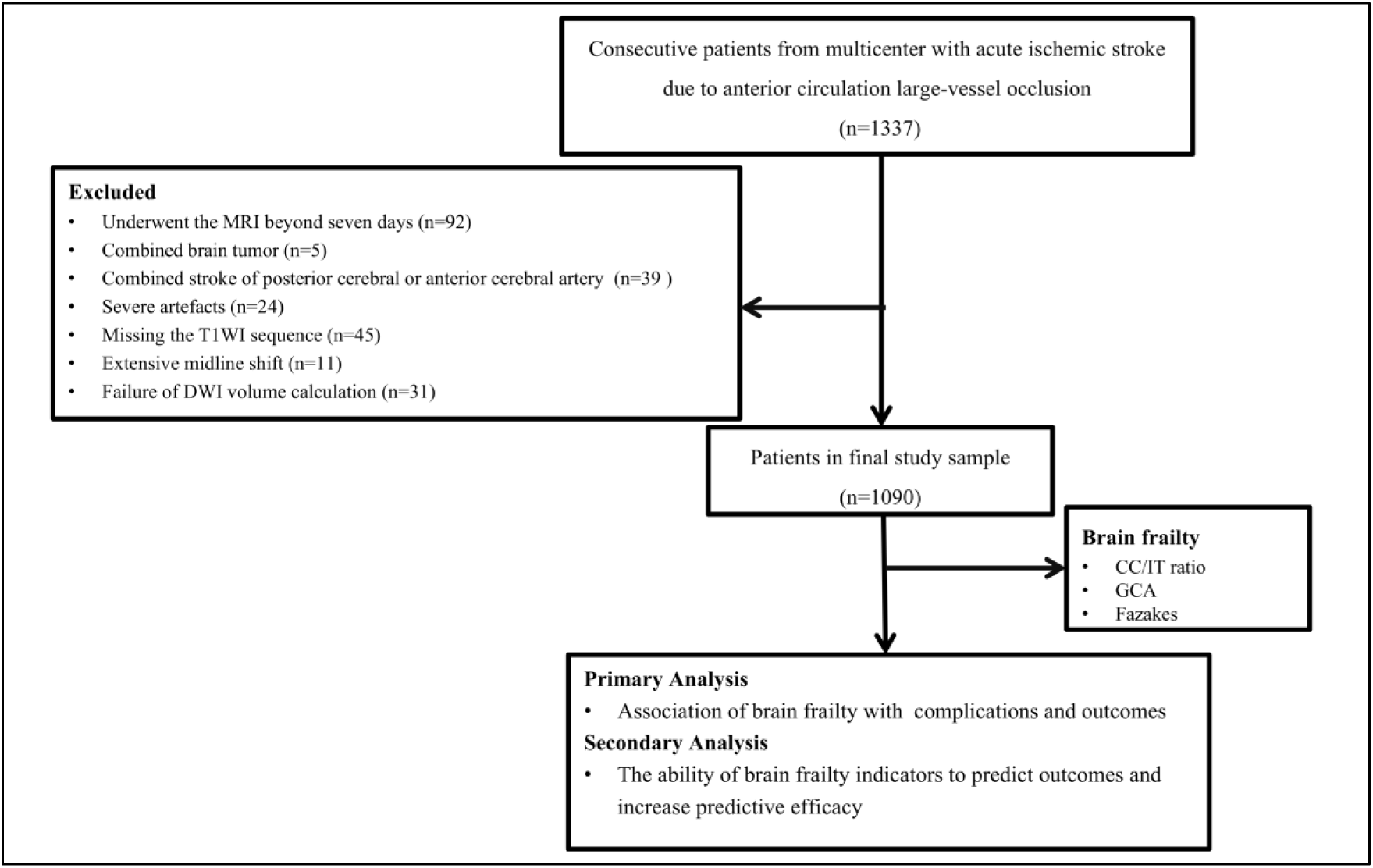
Patient selection flowchart. Abbreviation: CC/IT, intercaudate distance to inner table width;GCA, global cortical atrophy score

The median age of the remaining 1,090 participants was 64 (IQR, 55–73) years, and 754 (69.2%) were male. Endovascular treatment was given to 443 patients (40.6%), intravenous thrombolysis to 143 (13.1%), and medical therapy to 504 (46.3%). The median NIHSS score was 9 (IQR, 4.50–14.00), and the median infarct volume was 13.42 (IQR, 4.33–44.45) mL. Parenchymal hemorrhage was detected in 122 patients (11.2%), malignant cerebral edema in 42 (3.9%), and unfavorable 90-day clinical outcomes in 435 (39.9%).

A comparison of the data for patients with favorable and unfavorable clinical outcomes is shown in Table 1. Patients with unfavorable clinical outcomes had higher median age [67 (IQR, 58–75) vs. 62 (IQR, 52–70) years; *p* < 0.001], NIHSS score [13 (IQR, 8–17) vs. 7 (IQR, 3–12); *p* < 0.001], and infarct volume [26.08 (IQR, 7.44–101.13) vs. 10.12 (IQR, 3.51–26.37) mL; *p* < 0.001] than patients with favorable clinical outcomes. For brain atrophy indicators, patients with unfavorable clinical outcomes had more severe median subcortical atrophy [0.13 (IQR, 0.11–0.15) vs. 0.11 (IQR, 0.09–0.13); *P* < 0.001], severe cortical atrophy rate (8.51% vs. 1.98% ; *p* < 0.001), and Fazekas scores of 3 (12.64% vs. 3.97% ; *p* < 0.001) than those with favorable clinical outcomes.

**Table 1.** Clinical and imaging characteristics of the patient population

| Characteristic | All participants<br>(n = 1090) | Favorable outcome<br>(mRS 0–2, n = 655) | Unfavorable outcome<br>(mRS 3–6, n = 435) | P-value |
| --- | --- | --- | --- | --- |
| Age, y, median (IQR) | 64.00 (55.00-73.00) | 62.00 (52.00-70.00) | 67.00 (58.00-75.00) | <0.001 |
| Sex, male, n (%) | 754 (69.2%) | 479 (73.1%) | 275 (63.2%) | <0.001 |
| Baseline NIHSS, median (IQR) | 9.00 (4.50-14.00) | 7.00 (3.00-12.00) | 13.00 (8.00-17.00) | <0.001 |
| Infarction volume, median (IQR), mL | 13.42 (4.33-44.45) | 10.12 (3.51-26.37) | 26.08 (7.44-101.13) | <0.001 |
| **Laboratory data** |  |  |  |  |
| Blood glucose, median (IQR), mmol/L | 5.73 (4.88-7.33) | 5.40 (4.70-7.00) | 6.40 (5.18-8.08) | <0.001 |
| HbA1c, median (IQR), % | 5.90 (5.50-6.90) | 5.80 (5.45-7.00) | 5.90 (5.50-6.80) | 0.442 |
| Total cholesterol, median (IQR), mmol/L | 4.24 (3.62-4.98) | 4.21 (3.62-5.01) | 4.29 (3.62-4.91) | 0.587 |
| LDL, median (IQR), mmol/L | 2.48 (1.93-3.14) | 2.48 (1.94-3.13) | 2.50 (1.92-3.16) | 0.316 |
| HDL, median (IQR), mmol/L | 1.04 (0.88-1.24) | 1.03 (0.87-1.23) | 1.04 (0.89-1.27) | 0.581 |
| **Brain frailty** |  |  |  |  |
| CC/IT ratio, median (IQR) | 0.12 (0.10-0.14) | 0.11 (0.09-0.13) | 0.13 (0.11-0.15) | <0.001 |
| Global cortical atrophy score |  |  |  | <0.001 |
| 0-1 | 754 (69.2%) | 516 (78.8%) | 238 (54.7%) |  |
| 2 | 286 (26.2%) | 126 (19.2%) | 160 (36.8%) |  |
| 3 | 50 (4.6%) | 13 (2.0%) | 37 (8.5%) |  |
| Fazekas score |  |  |  | <0.001 |
| 0-1 | 706 (64.8%) | 488 (74.5%) | 218 (50.1%) |  |
| 2 | 303 (27.8%) | 141 (21.5%) | 162 (37.2%) |  |
| 3 | 81 (7.4%) | 26 (4.0%) | 55 (12.6%) |  |

| **Comorbidities** |  |  |  |  |
| --- | --- | --- | --- | --- |
| Hypertension, n (%) | 688 (63.2%) | 409 (62.4%) | 279 (64.3%) | 0.537 |
| Diabetes, n (%) | 229 (21.0%) | 136 (20.8%) | 93 (21.4%) | 0.792 |
| Atrial fibrillation, n (%) | 166 (15.3%) | 81 (12.4%) | 85 (19.6%) | 0.001 |
| Previous stroke, n (%) | 62 (5.7%) | 33 (5.0%) | 29 (6.7%) | 0.256 |
| **Treatment** |  |  |  | <0.001 |
| Medical therapy, n (%) | 504 (46.2%) | 342 (52.2%) | 162 (37.2%) |  |
| Endovascular therapy, n (%) | 443 (40.6%) | 250 (38.2%) | 193 (44.4%) |  |
| Intravenous thrombolysis, n (%) | 143 (13.1%) | 63 (9.6%) | 80 (18.4%) |  |

Abbreviations: CC/IT, intercaudate distance to inner table width; IQR, 25th–75th percentile; LDL, low-density lipoprotein; mRS, modified Rankin Scale; NIHSS, National Institutes of Health Stroke Scale; HbA1c, glycated hemoglobin; HDL, high-density lipoprotein

### Association of brain frailty with complications and outcomes

Multivariate logistic regression models adjusted for age, sex, NIHSS score, and infarct volume are shown in Table 2. For malignant cerebral edema, NIHSS score (OR, 1.13; 95% CI, 1.07–1.20; *p* < 0.001), blood glucose (OR, 1.30; 95% CI, 1.12–1.51; *p* < 0.001), and infarct volume (OR, 1.67; 95% CI, 1.24–2.25; *p* < 0.001) were independent risk factors, while subcortical atrophy, cortical atrophy, and leukoaraiosis were not (Table 2). For parenchymal hemorrhage, NIHSS score (OR, 1.08; 95% CI, 1.03–1.12; *p* < 0.001) and infarct volume (OR, 1.67; 95% CI, 1.38–2.02; *p* < 0.001) were independent risk factors, whereas none of the remaining variables were significantly associated (Table 2).

**Table 2.** Multivariate logistic regression models for complications and unfavorable outcomes

| Variable | Malignant cerebral edema |  | Parenchymal hemorrhage |  | Unfavorable outcome |  |
| --- | --- | --- | --- | --- | --- | --- |
|  | OR (95% CI) | P-value | OR (95% CI) | P-value | OR (95% CI) | P-value |
| Age, y | 1.01 (0.97, 1.05) | 0.624 | 1.02 (0.99, 1.05) | 0.121 | 0.99 (0.97, 1.00) | 0.078 |
| Sex, male | 1.18 (0.48, 2.85) | 0.720 | 0.96 (0.55, 1.69) | 0.895 | 0.48 (0.30, 0.76) | 0.002 |
| Baseline NIHSS | 1.13 (1.07, 1.20) | <0.001 | 1.08 (1.03, 1.12) | <0.001 | 1.17 (1.13, 1.22) | <0.001 |
| Infarction volume | 1.67 (1.24, 2.25) | <0.001 | 1.67 (1.38, 2.02) | <0.001 | 1.32 (1.15, 1.52) | 0.001 |
| Blood glucose | 1.30 (1.12, 1.51) | <0.001 | 1.06 (0.95, 1.18) | 0.312 | 1.15 (1.04, 1.26) | 0.005 |
| CC/IT ratio | 0.60 (0.15, 2.32) | 0.457 | 1.02 (0.42, 2.46) | 0.969 | 1.27 (1.18, 1.37) | <0.001 |
| Global cortical atrophy |  |  |  |  |  |  |
| 0-1 | Reference |  | Reference |  | Reference |  |
| 2 | 0.64 (0.23, 1.78) | 0.394 | 1.02 (0.53, 1.96) | 0.963 | 2.28 (1.36, 3.83) | 0.002 |
| 3 | 0.75 (0.08, 7.07) | 0.805 | 1.51 (0.46, 5.00) | 0.501 | 5.46 (1.71, 17.45) | 0.004 |
| Fazekas score |  |  |  |  |  |  |
| 0-1 | Reference |  | Reference |  | Reference |  |
| 2 | 0.81 (0.28, 2.33) | 0.701 | 0.63 (0.32, 1.22) | 0.173 | 2.09 (1.30, 3.35) | 0.002 |
| 3 | 0.87 (0.17, 4.53) | 0.868 | 1.04 (0.38, 2.87) | 0.936 | 4.68 (1.93, 11.31) | 0.001 |

Abbreviations: CC/IT, intercaudate distance to inner table width; mRS, modified Rankin Scale; NIHSS, National Institutes of Health Stroke Scale. Adjusted for age, sex, NIHSS score, and infarct volume.

For unfavorable clinical outcomes, NIHSS score (OR, 1.17; 95% CI, 1.13–1.22; *p* < 0.001), blood glucose (OR, 1.15; 95% CI, 1.04–1.26; *p* = 0.005), infarct volume (OR, 1.32; 95% CI, 1.15–1.52; *p* = 0.001), subcortical atrophy (OR, 1.27; 95% CI, 1.18–1.37; *p* < 0.001), severe cortical atrophy (OR, 5.46; 95% CI, 1.71–17.45, *p* = 0.004), and Fazekas score of 3 (OR, 4.68; 95% CI, 1.93–11.31; *p* = 0.001) were independent predictors of unfavorable clinical outcomes.

The cumulative distribution functions (CDFs) of brain frailty indicators associated with adverse clinical outcomes are shown in Figure 2. The CDFs illustrate that the cumulative probability of unfavorable clinical outcomes increases with the severity of subcortical atrophy (Figure 2A), cortical atrophy (Figure 2B), and leukoaraiosis (Figure 2C). The distribution of 90-day mRS scores for indicators of brain frailty is shown in Figure 3.

**Figure 2.**
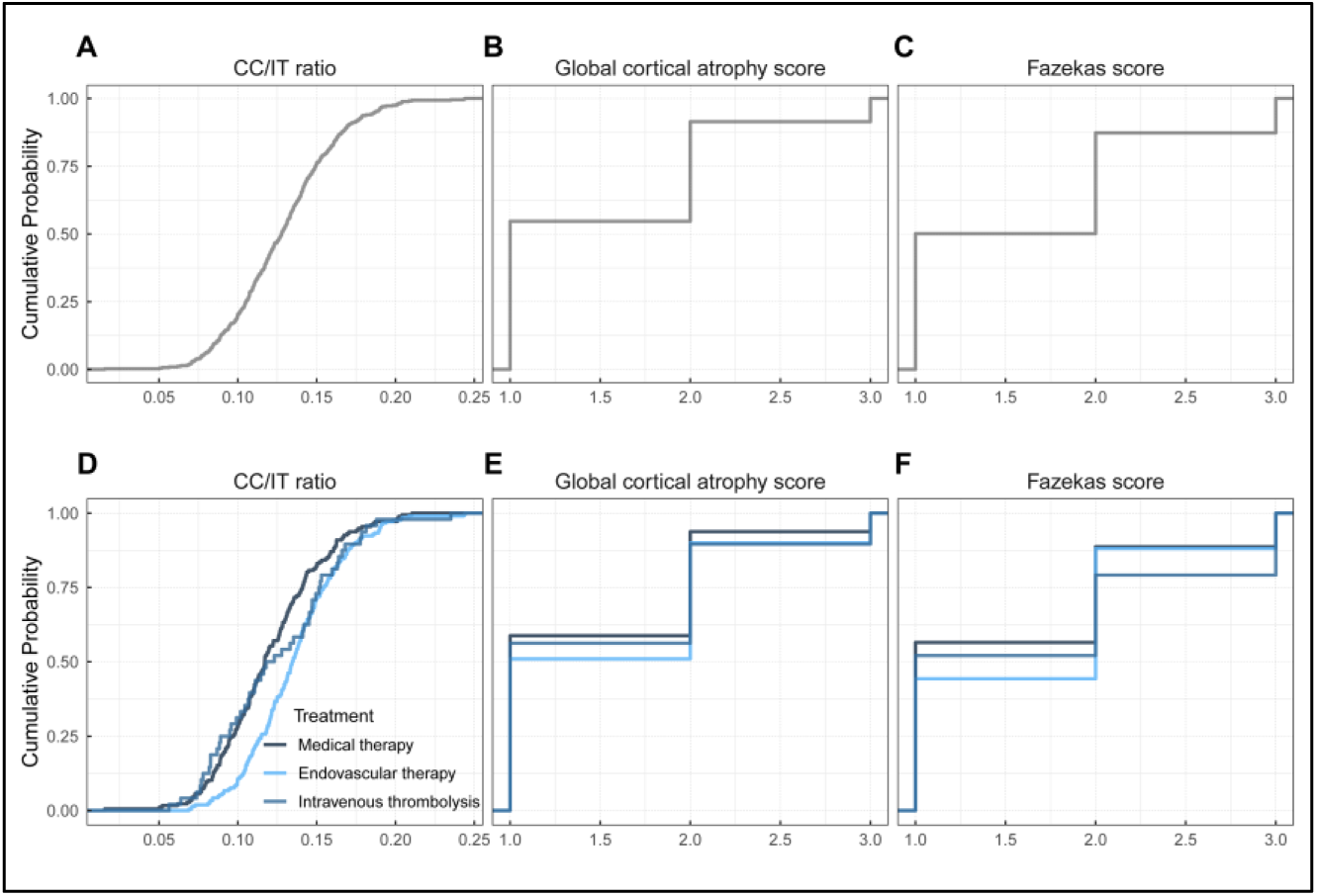
Cumulative distribution functions (CDFs) of brain frailty indicators with unfavorable clinical outcome (modified Rankin scale score 3–6). A–C In all patients with acute ischaemic stroke, the CDFs illustrate that the cumulative probability of unfavorable clinical outcomes increases with the severity of subcortical atrophy(CC/IT radio) (A), cortical atrophy (global cortical atrophy score) (B), and leukoaraiosis (Fazakes score)(C). D–F Subgroup analysis of different treatments to assess the cumulative probability distribution of subcortical atrophy (D), cortical atrophy (E) and leukoaraiosis (F) with unfavorable clinical outcome. Abbreviation: CC/IT, intercaudate distance to inner table width.

**Figure 3.**
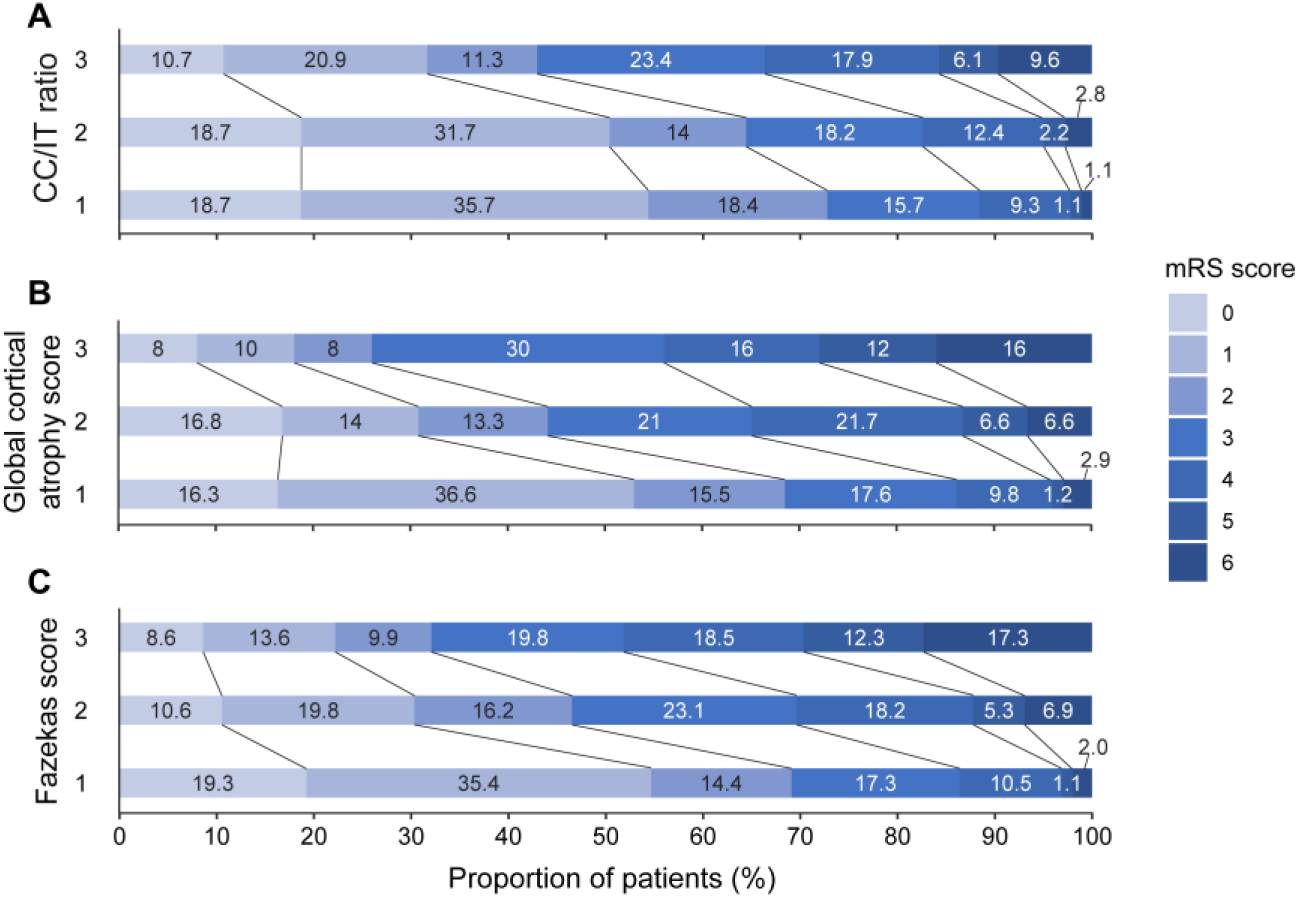
Distribution of 90-day mRS score for each indicator of brain frailty. A Distribution of 90-day mRS scores for subcortical atrophy (CC/IT ratio trisection); B Distribution of 90-day mRS scores for cortical atrophy (3-level score); C Distribution of 90-day mRS scores for leukoaraiosis (Fazekas score). Abbreviations: CC/IT, intercaudate distance to inner table width; mRS, modified Rankin Scale.

### ROC analysis for unfavorable clinical outcome

Univariable ROC analysis determined the AUCs for subcortical atrophy (0.655; 95% CI, 0.622–0.688), infarct volume (0.665; 95% CI, 0.632–0.699), blood glucose (0.623; 95% CI, 0.586–0.660), and NIHSS score (0.724; 95% CI, 0.694–0.754) (Supplementary Table S2). The diagnostic power of the NIHSS score was statistically better than that of infarct volume (DeLong test; *p* = 0.002) and subcortical atrophy (*p* = 0.003); infarct volume and subcortical atrophy had comparable predictive efficacy for unfavorable clinical outcomes (*p* = 0.609).

The unfavorable clinical outcome prediction model (model 1) developed with age, sex, NIHSS score, blood glucose, and infarct volume showed discriminatory power with an AUC of 0.762 (95% CI, 0.731–0.793), sensitivity of 64.0%, and specificity of 75.9%. The AUC of the model 1 increased significantly when brain atrophy indicators were included (Model 2; 0.822; 95% CI, 0.796–0.849; DeLong test; *p* < 0.001; Figure 4).

**Figure 4.**
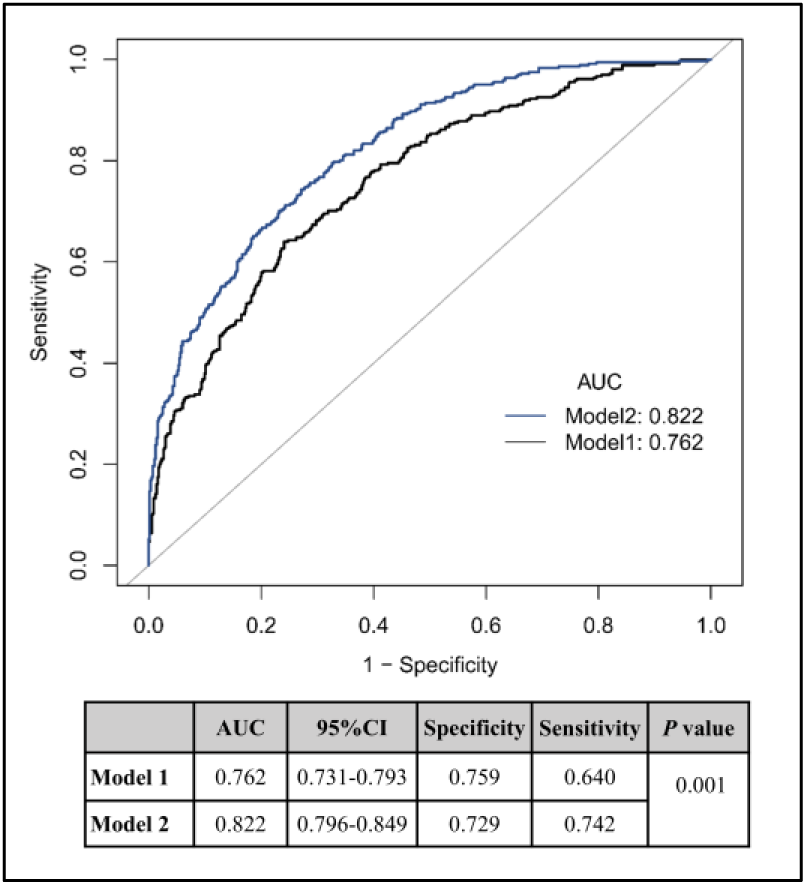
Receiver operating characteristic curves for predicting unfavorable clinical outcomes. Model 1: Age + Sex + NIHSS + Blood glucose + Infarction volume; Model 2: Model 1 + CC/IT ratio + GCA + Fazekas scores. Abbreviations: CC/IT, intercaudate distance to inner table width; GCA, global cortical atrophy score; NIHSS, National Institutes of Health Stroke Scale.

Subgroup analysis was conducted by treatment type, showing an increase in the AUC from Model 1 to Model 2 in all. For patients receiving endovascular treatment, the AUCs of Models 1 and 2 were 0.740 (95% CI, 0.686–0.793) and 0.872 (95% CI, 0.835–0.909), respectively. The respective AUCs for patients receiving medical therapy were 0.756 (95% CI, 0.710–0.802) and 0.812 (95% CI, 0.772–0.852). The respective AUCs for patients receiving intravenous thrombolysis were 0.849(95% CI, 0.781–0.917) and 0.888 (95% CI, 0.826–0.950) (Figure 5).

**Figure 5.**
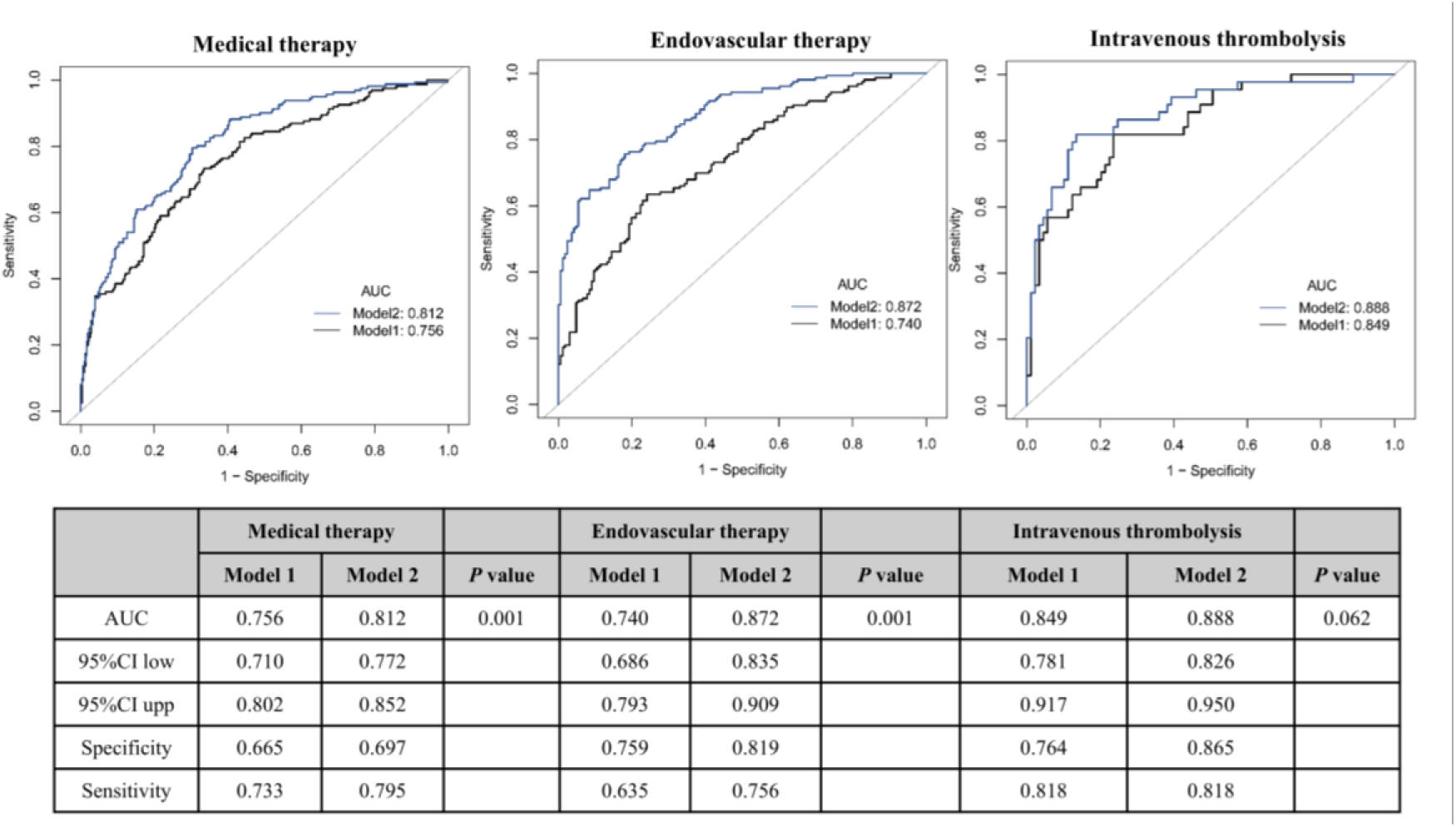
Receiver operating characteristic curves predicting unfavourable clinical outcomes (stratified analysis of treatment modalities). Model 1: Age+Sex+NIHSS+Blood glucose + Infarction volume; Model 2: Model 1+CC/IT ratio+GCA+Fazekas scores. Abbreviations: CC/IT, intercaudate distance to inner table width; GCA,global cortical atrophy score; mRS, modified Rankin Scale; NIHSS, National Institutes of Health Stroke Scale.

## DISCUSSION

This multicenter study identified 1,090 patients with AIS due to large-vessel occlusion of the anterior circulation and assessed the impact of brain atrophy indicators on acute complications and 90-day clinical outcomes. First, we confirm the prognostic values of subcortical atrophy, cortical atrophy, and leukoaraiosis, which were independently associated with unfavorable clinical outcomes. Second, quantitative measures of subcortical atrophy were comparable to infarct volume in predicting unfavorable clinical outcomes, and inclusion of brain atrophy indicators significantly improved the predictive power of the multivariate logistic regression models. In addition, no correlation was found between malignant cerebral edema or parenchymal hemorrhage secondary to AIS and brain atrophy. The results of this study highlight the importance of integrating brain atrophy into the clinical diagnostic predictions and therapeutic decision-making for patients with AIS and help understand the differential impact brain atrophy has on acute complications and long-term prognosis of stroke.

Brain atrophy reflects the resilience or vulnerability of the brain due to the accumulation of multiple chronic damages.^20^ The present study confirmed the important prognostic value of cortical atrophy, subcortical atrophy, and leukoaraiosis, each independently associated with unfavorable clinical outcomes. The ESCAPE-NA1 trial included patients with endovascular thrombolysis. Patients with cortical and subcortical atrophy detected by visual assessment of acute CT images had a less favorable AIS recovery and worse 90-day functional prognosis than patients without brain atrophy.^6^ A secondary analysis of that trial found that 85.1% of the association between age and 90-day functional outcomes after thrombectomy was mediated by brain frailty features, highlighting the importance of brain atrophy rather than chronological age alone in predicting stroke prognosis.^16^ In the International Stroke Trial (IST-3), which included patients with ischemic stroke who underwent intravenous thrombolysis, pre-existing brain CT signs (old infarcts, leukoaraiosis, and atrophy) were associated with decreased functional independence and increased symptomatic intracranial hemorrhage.^5^ The findings of our study, which included real-world patients with AIS receiving various treatment modalities, are consistent with the results described above, further demonstrating the importance of brain atrophy indicators for stroke prognosis. Furthermore, the brain atrophy indicators chosen for this study took into account clinical feasibility and reproducibility and are easily detectable on CT or MRI scans.^15^ The simple quantitative measurement of subcortical atrophy avoids the influence of the infarcted side of the brain, and visual assessment of cortical atrophy and leukoaraiosis is easy to perform without relying on any special software.

The prognostic predictive power of the model based on the remaining predictors of complications and unfavorable clinical outcomes found in our study, including age, NIHSS score, infarct volume, and blood glucose, improved significantly when combined with brain frailty indicators. Prognostic prediction models used in other studies relied on various variable combinations. For example, the THRIVE score included age, initial NIHSS score, and comorbidities.^9^ The SPAN-100 index summed the patient’s age and NIHSS score to predict clinical outcomes after intravenous and mechanical thrombolysis.^11^ The PRE score model combined the patient’s age, initial NIHSS score, and the Alberta Stroke Program Early Computed Tomography Score (ASPECTS) as continuous variables.^10^ The MRI-DRAGON score, which includes clinical (age, NIHSS score, glucose level, pre-stroke disability, and onset-to-treatment time) and radiological (DWI-ASPECTS, M1 occlusion) variables, had a high specificity for predicting the prognosis.^21^ None of these models included indicators of brain atrophy, and regardless of their complexity, their discriminative performance ranged between 0.73 and 0.83.^9,10,11,21^ A recent study showed that adding cortical atrophy and age-related white matter changes led to a small but significant improvement in machine learning-based clinical outcome prediction models, increasing the AUC from 0.763 to 0.775 .^22^ The present study used conventional variables and traditional logistic regression methods, achieving a significant increase in the AUC for predicting unfavorable clinical outcomes from 0.762 to 0.822 (*p* < 0.001) when including the brain atrophy indicators. The results remained stable in the treatment modality-based subgroup analyses.

Interesting differences in the correlation of brain atrophy with acute complications and functional recovery at 90 days were noted. The idea that brain atrophy might act as a buffer against acute stroke-induced brain swelling and occupancy effects has been previously suggested.^23,24^ A recent study investigated the effect of brain atrophy on the outcomes after endovascular treatment in 295 patients with AIS using visual scores and the Evans index (the ratio between the maximum widths of the frontal horns of the lateral ventricles and the maximal internal diameter of the skull at the same level). They found a positive association between the risk of futile reperfusion and atrophy; however, the Evans index does not account for underlying AIS-related edema or hemorrhage.^23^ A post hoc analysis of the ENCHANTED trial showed that brain atrophy was associated with lower early mortality in patients with stroke, possibly because brain atrophy made them more tolerant to brain swelling; however, the loss of neuronal plasticity and recovery also led to poorer functional prognosis.^24^ In our study, brain atrophy indicators did not significantly correlate with malignant brain edema or parenchymal hemorrhage, while the same indicators were risk factors for unfavorable 90-day functional outcomes. This difference indirectly reflects the different mechanisms through which brain atrophy affects acute complications and long-term prognosis. Clinical treatment decisions should take this difference into account.

The strengths of our study include the stability of the correlation between brain frailty and prognosis across various treatment modalities, and the difference between acute complications and 90-day prognosis in their correlation with brain frailty indicators. Moreover, the brain frailty indicators selected for this study are easy to measure and suitable for use in acute stroke settings and prospective studies. This study had some limitations: 1. Lack of finer segmentation and measurement of cortical and subcortical atrophy. In the absence of quantitative measurements of the leukoaraiosis volume, we used the Fazekas scale. 2. Some indicators of cerebral small vessel disease, such as microhemorrhages, lacunae, and enlarged perivascular spaces, were not assessed, given the commonality of the indicators between CT and MRI assessments. 3. The small number of patients receiving IV thrombolysis may have biased the study finding. 4. The model in this study obtained the prediction results for unfavorable clinical outcome through multivariate logistic regression analysis; because of the lack of cross-validation, it is possible that the results of the model are overly optimistic.

In conclusion, the present study suggests that brain frailty indices are strong independent prognostic predictors of AIS due to large-vessel occlusion; however, they are not associated with acute complications. These findings have important implications for guiding patients’ acute treatment and functional recovery. Brain frailty indicators significantly enhanced the prediction of patient prognosis. The selection of simple, easy-to-measure indicators of brain frailty that can be easily integrated into existing predictive models could support prospective therapeutic decision-making.

## Data Availability

The data that support the findings of this study are available on request from the corresponding author

## Acknowledgments

None

## Non-standard Abbreviations and Acronyms

AIS: acute ischemic stroke
AUC: area under the receiver operating characteristic curve
CDFs: cumulative distribution functions
DWI: diffusion-weighted imaging
FLAIR: fluid-enhanced inversion recovery
mRS: modified Rankin Scale
NIHSS: National Institutes of Health Stroke Scale
ROC: receptor operating characteristic
T1WI: T1-weighted imaging
T2WI: T2-weighted imaging

